# Classification of Fetal State through the application of Machine Learning techniques on Cardiotocography records: Towards Real World Application

**DOI:** 10.1101/2021.06.03.21255808

**Authors:** Andrew Maranhão Ventura Dadario, Christian Espinoza, Wellington Araújo Nogueira

## Abstract

**Objective:** Anticipating fetal risk is a major factor in reducing child and maternal mortality and suffering. In this context cardiotocography (CTG) is a low cost, well established procedure that has been around for decades, despite lacking consensus regarding its impact on outcomes.

Machine learning emerged as an option for automatic classification of CTG records, as previous studies showed expert level results, but often came at the price of reduced generalization potential.

With that in mind, the present study sought to improve statistical rigor of evaluation towards real world application.

**Materials and Methods:** In this study, a dataset of 2126 CTG recordings labeled as normal, suspect or pathological by the consensus of three expert obstetricians was used to create a baseline random forest model.

This was followed by creating a lightgbm model tuned using gaussian process regression and post processed using cross validation ensembling.

Performance was assessed using the area under the precision-recall curve (AUPRC) metric over 100 experiment executions, each using a testing set comprised of 30% of data stratified by the class label.

**Results:** The best model was a cross validation ensemble of lightgbm models that yielded 95.82% AUPRC.

**Conclusions:** The model is shown to produce consistent expert level performance at a less than negligible cost. At an estimated 0.78 USD per million predictions the model can generate value in settings with CTG qualified personnel and all the more in their absence.

## 1. Introduction

Direct information regarding the fetus well-being is not trivially acquired during pregnancy, and key information such as the fetal heart rate (FHR) is crucial for anticipating fetal risks in both antepartum as well as intrapartum. [12]

In this context cardiotocography (CTG) is a well-established, routine procedure that has been used since the end of the 1960s for monitoring the fetal heart rate and uterine contractions (UC) signals during pregnancy and delivery. [12]

The fetal heart rate itself is used for investigating the oxygen supply for the fetus, as hypoxia during labor can lead to death and long-term disabilities. [7]

The interpretation of the CTG signals is supported by guidelines developed by institutions such as International Federation of Gynecology and Obstetrics (FIGO) and the Institute of Child Health and Human Development (NICHD). [3]

While FHR and UC signals are the primary objective of CTG, the guidelines extend the definition of observations to include features that describe these signals, such as acceleration, deceleration, and variability. [16]

Despite the existence of these guidelines, the CTG exam is still prone to subjectivity and there is no universal consensus regarding its interpretation. This subjectivity extends to measuring its outcomes, which 40 or so years after its implementation, still retains significant variance. [2]

CTG is most commonly applied in high-risk pregnancies and is not recommended by the World Health Organization (WHO) for healthy pregnant women undergoing spontaneous labor. [20]

## 2. Related Work

Studies regarding the impact on outcomes of the CTG exam have considerable variability: sensitivity ranging from 2 to 100% and specificity between 37 and 100% [2].

At the heart of such variance lies the opportunity of improving consistency through the application of a machine-based model, which would remove inter-observer variation.

In the past studies exploring machine learning for automatic CTG classification, authors often favored theoretical performance over practical applications, as decisions such as excluding the suspect class [9] [15] [17], single run experiments [9] [15] [17] [21] and small sized testing sets [9] [17] [21] constrain the potential generalization of any given model.

The present study sought to improve the statistical rigor of previous work done with machine learning applied to CTG in order to bring the results one step closer to real world application.

## 3. Materials and Methods

### 3.1 Dataset

The dataset used in this study contains 2126 fetal cardiotocograms represented in 21 features belonging to 3 different classes: normal (n = 1655), suspect (n = 295) and pathological (n = 196). [1]

The class labels were given by the consensus of three expert obstetricians, using FIGO as its guideline for interpretation. [3]

The features were created by SisPorto 2.0 software [1], which applies pattern recognition to digital CTG signals yielding the features described in the table 1 below.

**Table 1.** Description of dataset variables. [3]

| Column Name | Description |
| --- | --- |
| LB | FHR baseline (beats per minute) |
| AC | # of accelerations per second |
| FM | # of fetal movements per second |
| UC | # of uterine contractions per second |
| DL | # of light decelerations per second |
| DS | # of severe decelerations per second |
| DP | # of prolonged decelerations per second |
| ASTV | Percentage of time with abnormal short-term variability |
| MSTV | Mean value of short-term variability |
| ALTV | Percentage of time with abnormal long-term variability |
| MLTV | Mean value of long-term variability |
| Width | Width of FHR histogram |
| Min | Minimum of FHR histogram |
| Max | Maximum of FHR histogram |
| Nmax | # of histogram peaks |
| Nzeros | # of histogram zeros |
| Mode | Histogram mode |
| Mean | Histogram mean |
| Median | Histogram median |
| Variance | Histogram variance |
| Tendency | Histogram tendency |
| NSP | Fetal state class code (N=normal; S=suspect; P=pathologic) |

### 3.2 Metrics

The primary metric used to measure performance in this study is the **area under the precision-recall curve**. The reasoning behind this is trifold: the metric is representative of performance amidst class imbalance, the metric allows practical decisions (i.e., favoring recall for screening purposes or precision for resource allocation) as well as being conceptually familiar to professionals that underwent nursery or medical school. The AUPRC can be defined in the equation below [14], where *p* and *r* denote precision and recall respectively:

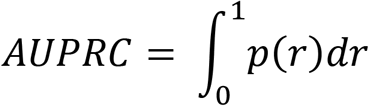

Secondary metrics were also made available, including accuracy, precision, recall, f1-score and area under the receiver operator characteristic curve (AUROC).

AUPRC was chosen in favor of AUROC as the ROC curve can be misleading in the face of class imbalance, as few examples of the minority class diminish the trustworthiness of the measured performance [5].

Logloss was used as a loss function for training classifiers as well as the minimization criteria during hyperparameter optimization.

### 3.3 Evaluation Method

In order to ensure the reliability of proposed techniques, all experiments were repeated 100 times (n = 100) and the reported metrics represent the median of experiment runs.

In every experiment run, the testing set was composed by 30% of data using the target classes as stratification criteria.

During hyperparameter optimization, models were trained under k-fold cross validation using k = 4 on training data.

### 3.4 Machine Learning Models and Hyperparameter Optimization

In order to establish the baseline performance level, a RandomForest model [6] was conceived. The reasoning behind this choice is due to the low variance coupled with good bias levels as well as the synergy between this framework and the final candidate, a LightGBM model [13].

Tree-based algorithms rely on similar assumptions and representations, therefore its performance can be consistently compared whilst not incurring any extra overhead for preprocessing.

The parameters for the baseline model were not tuned, rather, they were chosen for the main purpose of lower variance, as to establish a consistent baseline for performance while also minimizing bias whenever possible.

The table 2 reports the parameters and constants used in the baseline model.

**Table 2.**
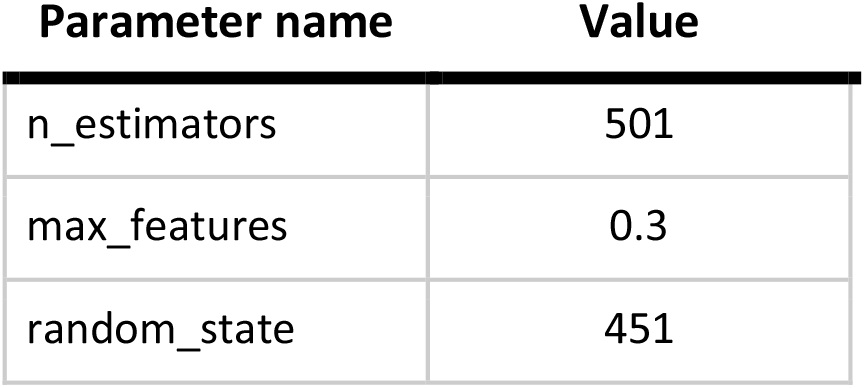
Baseline random forest model parameters.

**Table 3.** LightGBM parameters obtained through bayesian optimization.

| Parameter name | Value |
| --- | --- |
| learning_rate | 0.034086444079214386 |
| n_estimators | 300 |
| num_leaves | 31 |
| max_depth | 11 |
| max_bin | 356 |
| bagging_freq | 8 |
| bagging_fraction | 0.6433384789192684 |
| feature_fraction | 0.700623286108986 |
| min_child_samples | 7 |
| min_split_gain | 0.0 |
| boosting_type | gbdt |
| bagging_seed | 42 |
| random_state | 451 |

After the baseline model, a lightgbm model was conceived through bayesian optimization using a gaussian process regression mapping the logloss of the model (calculated on k-fold cross validation with k = 4) to the parameters in the search space.

The optimization procedure had 30 random starts followed by 70 rounds of refinement leading to a lightgbm classifier with the following parameters:

### 3.5 Post Processing

Following the results of hyperparameter optimization, the resulting LightGBM model was subjected to k-fold cross validation ensembling (CVE) wherein the model is trained multiple times (k = 4) on different subsets of the training set and its final predictions are subsequently averaged. This process managed to reduce both bias and variance, as shown in the next section.

## 4. Results

The results are summarized in table 6 and charts 1 and 2 depict the performance of the best model. A more detailed table of experiment results per model is available on annex 1, 2 and 3.

| Parameter name | AUPRC | Accuracy | AUROC | F1-Score | Recall | Precision |
| --- | --- | --- | --- | --- | --- | --- |
| Random Forest Baseline | 0.9559 | 0.9436 | 0.9868 | 0.8991 | 0.8831 | 0.9182 |
| LightGBM | 0.9577 | 0.9483 | 0.9877 | 0.9084 | 0.8932 | 0.9261 |
| **CVE LightGBM** | **0.9582** | **0.9514** | **0.9880** | **0.9128** | **0.8986** | **0.9286** |

**Chart 1:**
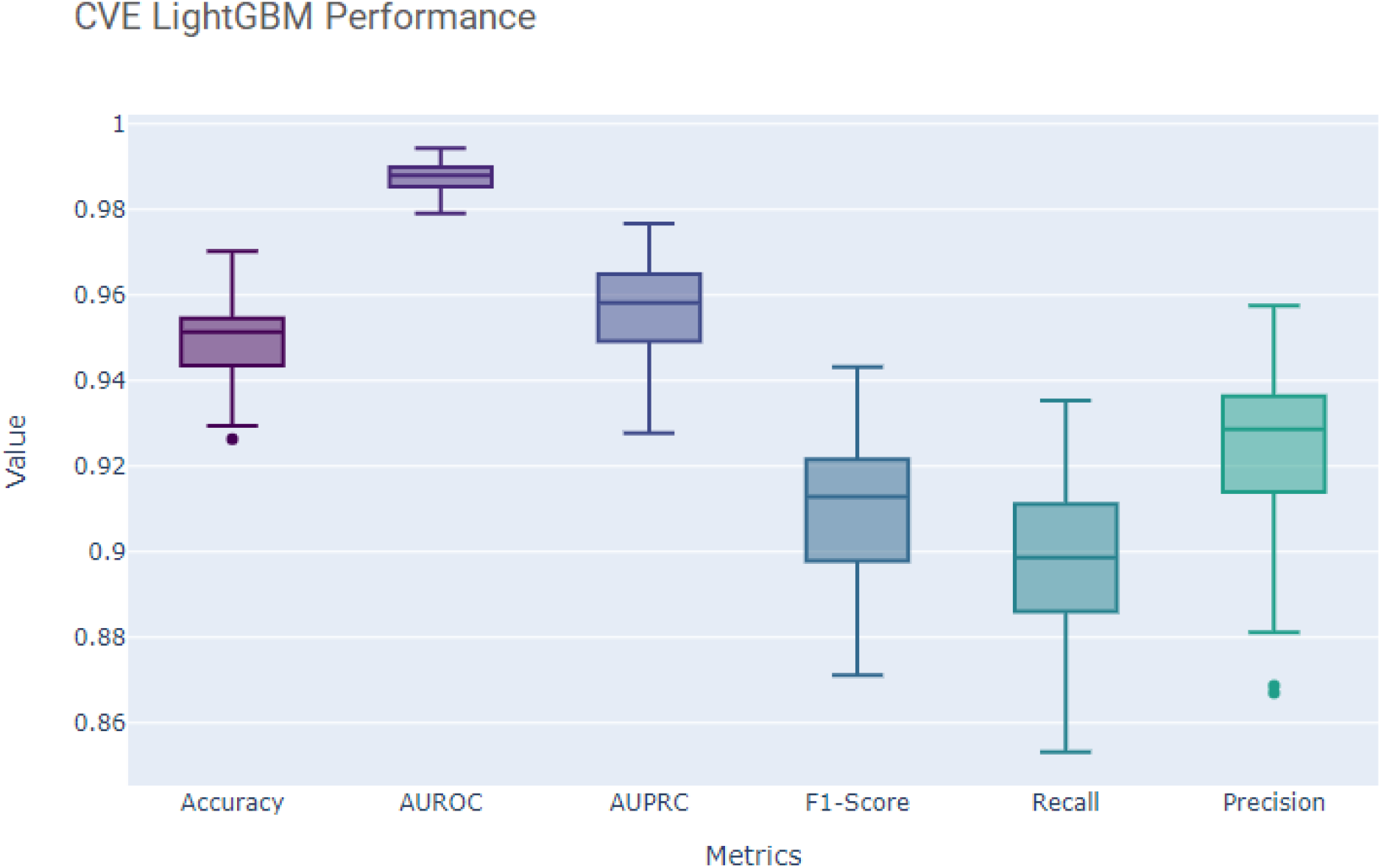
Performance of CVE LightGBM

**Chart 2:**
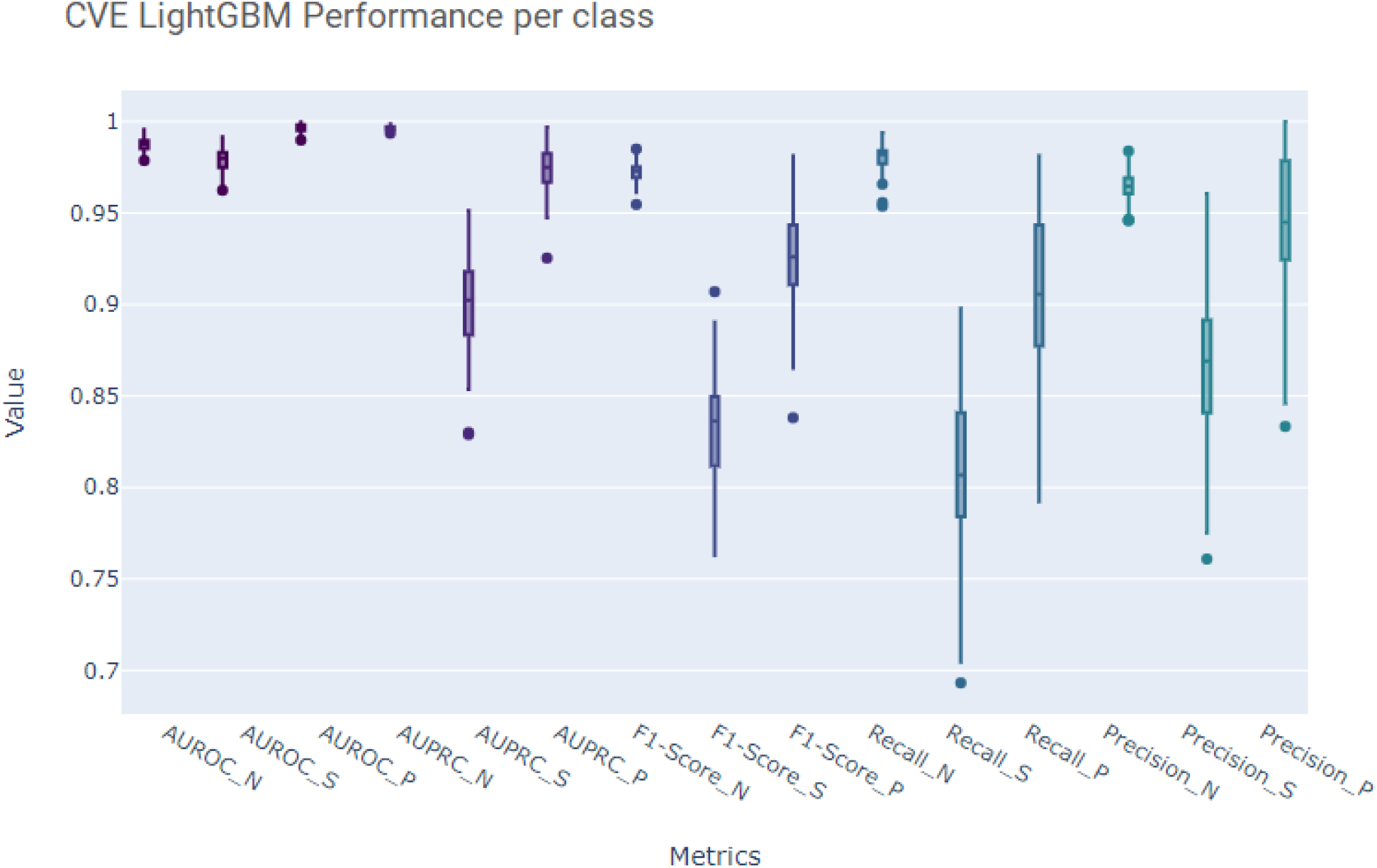
Performance of CVE LightGBM measured by class Note: N denotes normal, S denotes suspect and P, pathological.

As part of the results, a cost estimate for the model was made in order to further support real world deployment and healthcare value assessment.

From an estimate of 140 million yearly births [19] worldwide, roughly translating to 12 million births/month, and 300 milliseconds execution time, the lightgbm model would cost 0.78 USD for every million predictions or 9.33 USD per month for covering all births in that given period.

## 5. Discussion

The suspect class was shown to be the most difficult to predict, likely due to the fact that there are less observations when compared to the normal class as well as being the in-between of the other classes as well.

While it would certainly increase the computational overhead, using the raw signal could yield better results, as theoretically the bayes optimal error is diminished when using aggregations like it was done by SisPorto 2.0.

The cost estimate does not factor indirect costs that are facility specific, such as IT, data infrastructure required to support the model, and it drastically overestimates the amount of predictions required, as not all labor occurrences would need a CTG exam to begin with.

In order to further approach this model to a real-world setting, we recommend exploring the effects of manufacturer, age and ethnic group [4] in order to ensure that the model retains performance levels amidst populational and hardware variance.

## 6. Conclusion

The models created in the course of this study showed good and consistent levels of performance. Lightgbm with bayesian optimization proved very useful in pushing the baseline, as did cross validation ensemble which introduced a small but welcome performance gain.

As the impact on outcomes of CTG remains unclear, a machine model could improve measurements by reducing subjectivity.

The low-cost structure combined with the fact that CTG is a widespread procedure makes it a great candidate for real world experimentation.

The cost overhead added by the AI model is easily overshadowed by the potential efficiency gains in domains with CTG qualified professionals and even more so for resource poor environments where the exam would be otherwise unavailable.

## Data Availability

The dataset used in this study is publicly available.

https://archive.ics.uci.edu/ml/datasets/cardiotocography

## 7. Acknowledgements

We would like to thank Biraja Machado, Edson Amaro, Adriano Pereira and Wellington Lucena for the inspiration and support. We also express our gratitude to Kaggle for providing free computational resources.

## 9. Annex

**Annex 1.** Random Forest baseline experiment results

|  | **Accuracy** | **AUROC** | **AUPRC** | **F1-Score** | **Recall** | **Precision** |
| --- | --- | --- | --- | --- | --- | --- |
| Mean | 0,9432 | 0,9865 | 0,9541 | 0,8967 | 0,8804 | 0,9172 |
| Std | 0,0082 | 0,0031 | 0,0094 | 0,0157 | 0,0185 | 0,0185 |
| Minimum | 0,9232 | 0,9787 | 0,9311 | 0,8581 | 0,8395 | 0,8559 |
| 25% | 0,9373 | 0,9843 | 0,9471 | 0,8868 | 0,8667 | 0,9070 |
| Median | 0,9436 | 0,9868 | 0,9559 | 0,8991 | 0,8831 | 0,9182 |
| 75% | 0,9487 | 0,9889 | 0,9610 | 0,9076 | 0,8938 | 0,9320 |
| Maximum | 0,9592 | 0,9926 | 0,9711 | 0,9263 | 0,9182 | 0,9521 |

**Annex 2.** LightGBM experiment results

Annex 1. Random Forest baseline experiment results
|  | **Accuracy** | **AUROC** | **AUPRC** | **F1-Score** | **Recall** | **Precision** |
| --- | --- | --- | --- | --- | --- | --- |
| Mean | 0,9484 | 0,9873 | 0,9561 | 0,9074 | 0,8939 | 0,9237 |
| Std | 0,0083 | 0,0033 | 0,0112 | 0,0169 | 0,0196 | 0,0187 |
| Minimum | 0,9248 | 0,9775 | 0,9255 | 0,8652 | 0,8452 | 0,8680 |
| 25% | 0,9436 | 0,9852 | 0,9491 | 0,8975 | 0,8825 | 0,9133 |
| Median | 0,9483 | 0,9877 | 0,9577 | 0,9084 | 0,8932 | 0,9261 |
| 75% | 0,9545 | 0,9897 | 0,9642 | 0,9186 | 0,9092 | 0,9358 |
| Maximum | 0,9671 | 0,9948 | 0,9781 | 0,9456 | 0,9360 | 0,9601 |

**Annex 3.** Cross validation ensemble LightGBM experiment results

Annex 2. LightGBM experiment results
|  | **Accuracy** | **AUROC** | **AUPRC** | **F1-Score** | **Recall** | **Precision** |
| --- | --- | --- | --- | --- | --- | --- |
| Mean | 0,9498 | 0,9877 | 0,9573 | 0,9097 | 0,8968 | 0,9251 |
| Std | 0,0085 | 0,0032 | 0,0107 | 0,0169 | 0,0196 | 0,0182 |
| Minimum | 0,9263 | 0,9791 | 0,9277 | 0,8712 | 0,8532 | 0,8670 |
| 25% | 0,9436 | 0,9853 | 0,9494 | 0,8979 | 0,8861 | 0,9141 |
| Median | 0,9514 | 0,9880 | 0,9582 | 0,9128 | 0,8986 | 0,9286 |
| 75% | 0,9545 | 0,9898 | 0,9647 | 0,9215 | 0,9110 | 0,9362 |
| Maximum | 0,9702 | 0,9943 | 0,9767 | 0,9432 | 0,9353 | 0,9575 |

